# The Impact of Physician-Patient Gender Match on Healthcare Quality: An Experiment in China

**DOI:** 10.1101/2023.10.03.23296202

**Authors:** Yafei Si, Gang Chen, Min Su, Zhongliang Zhou, Winnie Yip, Xi Chen

## Abstract

Despite growing evidence of gender disparities in healthcare utilization and health outcomes, there is a lack of understanding of what may drive such differences. Designing and implementing an experiment using the standardized patients’ approach, we present novel evidence on the impact of physician-patient gender match on healthcare quality in a primary care setting in China. We find that, compared with female physicians treating female patients, the combination of female physicians treating male patients resulted in a 23.0 percentage-point increase in correct diagnosis and a 19.4 percentage-point increase in correct drug prescriptions. Despite these substantial gains in healthcare quality, there was no significant increase in medical costs and time investment. Our analyses suggest that the gains in healthcare quality were mainly attributed to better physician-patient communications, but not the presence of more clinical information. This paper has policy implications in that improving patient centeredness and incentivizing physicians’ efforts in consultation (as opposed to treatment) can lead to significant gains in the quality of healthcare with modest costs, while reducing gender differences in care.

## Introduction

Gender is one of the first characteristics we notice in others. This acute awareness of gender can be problematic, because our gender-related thinking, feeling, and behavioral responses to others can lead to undesirable gender disparities. For example, women often fare worse than men in equitable pay (Cohen and Huffman, 2007), ascension to leadership positions (Cohen et al., 1998), educational outcomes (Carlana, 2019; Carrell et al., 2010), scholarly peer-review processes (Card et al., 2020), and legal dispute resolution (Knepper, 2018). In the medical setting, prevalent gender disparities that disadvantage female patients are reported in access to critical health interventions (Hoffmann and Tarzian, 2001; Milcent et al., 2007), potentially life-saving treatments (Clarke et al., 1994; Hamberg, 2008), and health outcomes (Shannon et al., 2019). However, little is known about what drives these disparities and what policy interventions may affect them. To address this gap, we investigate the role of physician-patient gender match in explaining gender-related disparities in healthcare quality.

Healthcare quality determines the extent to which health services for individuals and populations increase the likelihood of desired health outcomes (WHO, 2022). Existing findings suggest significant gaps between what physicians know how to do and what they do in practice (Leonard and Masatu, 2010; Mohanan et al., 2015). The low quality of healthcare attributed to “know-do” gaps is of general concern in developing countries (Brownlee et al., 2017; Das et al., 2012; Su et al., 2021; Sylvia et al., 2015), especially for female patients. In practice, patients often know little about the quality of care they receive (Arrow, 1963; Dulleck and Kerschbamer, 2006), while their decisions on treatments heavily rely on physicians’ advice. Growing evidence from developed countries suggests that physicians do not practice solely based on science but vary their behaviors and choices depending on values, beliefs, race, and gender (Greenwood et al., 2018; Gross et al., 2008; Malhotra et al., 2017). It is challenging to isolate gender effects from other unobservable factors (i.e., modified patient behavior, biological factors, and latent health status) using observational study designs (Cabral and Dillender, 2021; Daniels et al., 2019; Wallis et al., 2021; Weisse et al., 2005). For instance, patients have preferences regarding physician gender (Janssen and Lagro-Janssen, 2012), which results in selection biases. Therefore, how physician-patient gender match determines healthcare quality is poorly understood, partly due to methodological challenges.

Designing and implementing an experiment using the standardized patients (SPs) method, we assess the impact of physician-patient gender match on healthcare quality. SPs are well-coached actors who are trained to present symptoms of an illness to physicians in a standardized, unvarying manner and to then record in detail the interactions between them (Wiseman et al., 2019). SPs present their initial symptoms and respond to physicians’ inquiries consistently across all interactions, and thus unobservable factors from the patient side that may confound any identified relationship between physician-patient gender match and healthcare quality can be mitigated. The SP method has been widely used in medical education in developed countries and is increasingly used to evaluate healthcare quality in developing countries as well (Das et al., 2016b). We leverage the exogenous variation of assigning SPs to randomly pair with primary care physicians, due to the walk-in nature of primary care in our setting, to evaluate the impact of physician-patient gender match.

## Methods

The study was conducted in Xi’an, China between August 17–28, 2017 and between July 30–August 10, 2018. We recruited 18 SPs (person-years) from the local communities, of whom 15 were female and 3 were male. The SPs participated in rigorous training before seeing physicians at community health centers (CHCs). During the audit study, each SP was offered compensation of 200 Chinese Yuan (CNY) per day (the average local daily earning rate) and an additional bonus (1,000 CNY) if they participated in the whole study (and all SPs did).

The SPs portrayed two gender-neutral diseases, unstable angina and asthma, which were highly prevalent in the study region. The two conditions were selected because they allowed SPs’ opening statements to appear consistent with multiple underlying illnesses, meaning that further inquiries would be required by the physician to produce unambiguous and correct diagnoses and treatment (King et al., 2019). The scripts for the two conditions were validated in both India and China (Das et al., 2012; Sylvia et al., 2015), and the risk of invasive medical tests being necessary was low.

During the visits, SPs answered physicians’ inquiries, accepted all non-invasive medical tests, and paid for the consultation, medical tests, and all medications. We required SPs to report their detailed interactions with physicians by administering a structured questionnaire immediately after they left the CHC. We then further checked SPs’ responses using an audio recording to ensure accuracy. In total, we collected 492 interactions between 169 physicians and 18 SPs. The details regarding data collection have been published (Si et al., 2023).

All 63 CHCs in Xi’an, China granted approval for the study to be conducted in their facilities. Written consent forms were obtained from CHCs and physicians three months before the SPs’ visits, but physicians were not aware of the diseases to be tested. The study was approved by the Ethics Committee of Xi’an Jiaotong University Health Science Center (approval number: 2015-406); this committee also permitted the recording of interactions between physicians and SPs using hidden audio devices.

### Healthcare Quality

We focus on one essential aspect of quality: the degree to which patients receive timely and accurate diagnoses and evidence-based treatment for their conditions without facing financial hardship (Das et al., 2018). We use four metrics to measure healthcare quality.

1. *Consultation length* refers to how long (in minutes) the physician-patient communication lasts and is used as a proxy for provider effort (Das et al., 2016b).
2. *Medical costs* include the fees incurred for consultation, medical tests, and drugs prescribed.
3. *Correct diagnosis:* We use a binary variable to classify diagnoses as “correct” or “other” (i.e., partially correct or incorrect—see *Supplement 1*). SPs were instructed to consult physicians directly at the end of their visits if a diagnosis had not been provided.
4. *Correct drug prescription:* We use another binary variable to classify drugs as “correct” or “other” (i.e., unnecessary or harmful—see *Supplement 2*) (Das et al., 2016b; Sylvia et al., 2017).

### The Empirical Model

Our econometric specification is very straightforward:

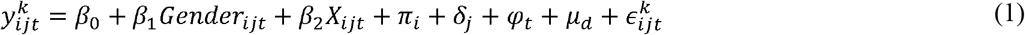

where 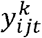 represents quality metric k in the CHC *i* and district *j* on day *t. Gender*_*ijt*_ is specified to control for four pairs of physician–patient gender match using the pair of female physicians treating female SPs as a reference group. *X*_*ijt*_ is a set of observable demographic factors of physicians. *π*_*i*_ indicates CHC fixed effects, *δ*_*j*_ indicates district fixed effects, and *µ*_*d*_ indicates disease fixed effects. *φ*_*t*_ indicates month, day of the week, and year fixed effects. 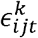 is the error term. Linear regression models were estimated. Robust standard errors were clustered at the CHC level.

## Results

Female physicians accounted for 54.47% of physician–patient interactions, while female SPs accounted for 83.54% of interactions. In terms of physician–patient pairs, female physicians treating female SPs accounted for 46.14% of all interactions, male physicians treating female SPs accounted for 37.40% of all interactions, female physicians treating male SPs accounted for 8.33% of all interactions, and male physicians treating male SPs accounted for 8.13% of all interactions. The observable CHC and physician characteristics were statistically balanced across the four pairs of physician–patient gender match *(Table S1)*. However, female physicians tended to be younger than male physicians in our setting, and thus the effects of physicians’ age were adjusted for in the regression analysis.

We summarize the quality metrics per the four pairs of physician–patient gender match. Female physicians treating male SPs was significantly associated with a higher probability of prescribing correct drugs but higher medical costs (*Figure 1*) compared with the other pairs. We do not find any significant difference in the consultation length, or the probability of correct diagnosis. We further estimate regression models to examine the impact of physician–patient gender match on healthcare quality. In doing this, we find that, compared with female physicians treating female SPs, female physicians treating male SPs resulted in a significant 23.0 percentage-point increase in correct diagnosis and a 19.4 percentage-point increase in correct drug prescriptions (*Figure 2*). However, we find no significant change in medical costs or the length of consultation.

**Figure 1.**
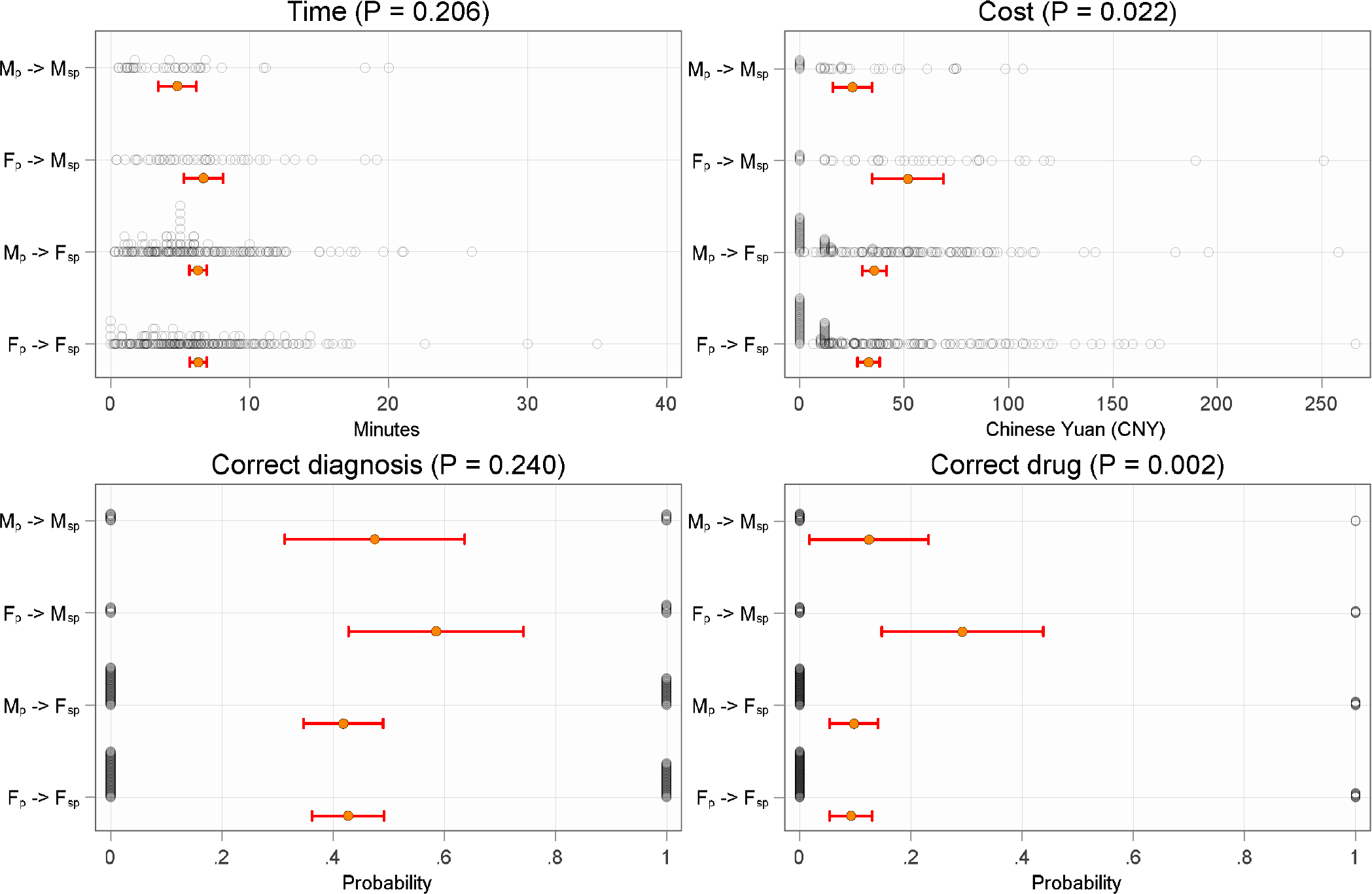
Quality metrics over physician–patient gender match Note: The figure reports mean and 95% confidence intervals for all quality metrics, including the distribution of all observations. The pair of *F*_*p*_ -> *F* _*sp*_ denotes female physicians treating female patients (SPs) and others likewise. CNY denotes Chinese yuan (exchange rate, 6.37 CNY ≈ 1 US dollar). The statistical differences were analyzed using the chi-square test for binary variables and analysis of variance for continuous variables. The details are reported in *Table S2*.

**Figure 2.**
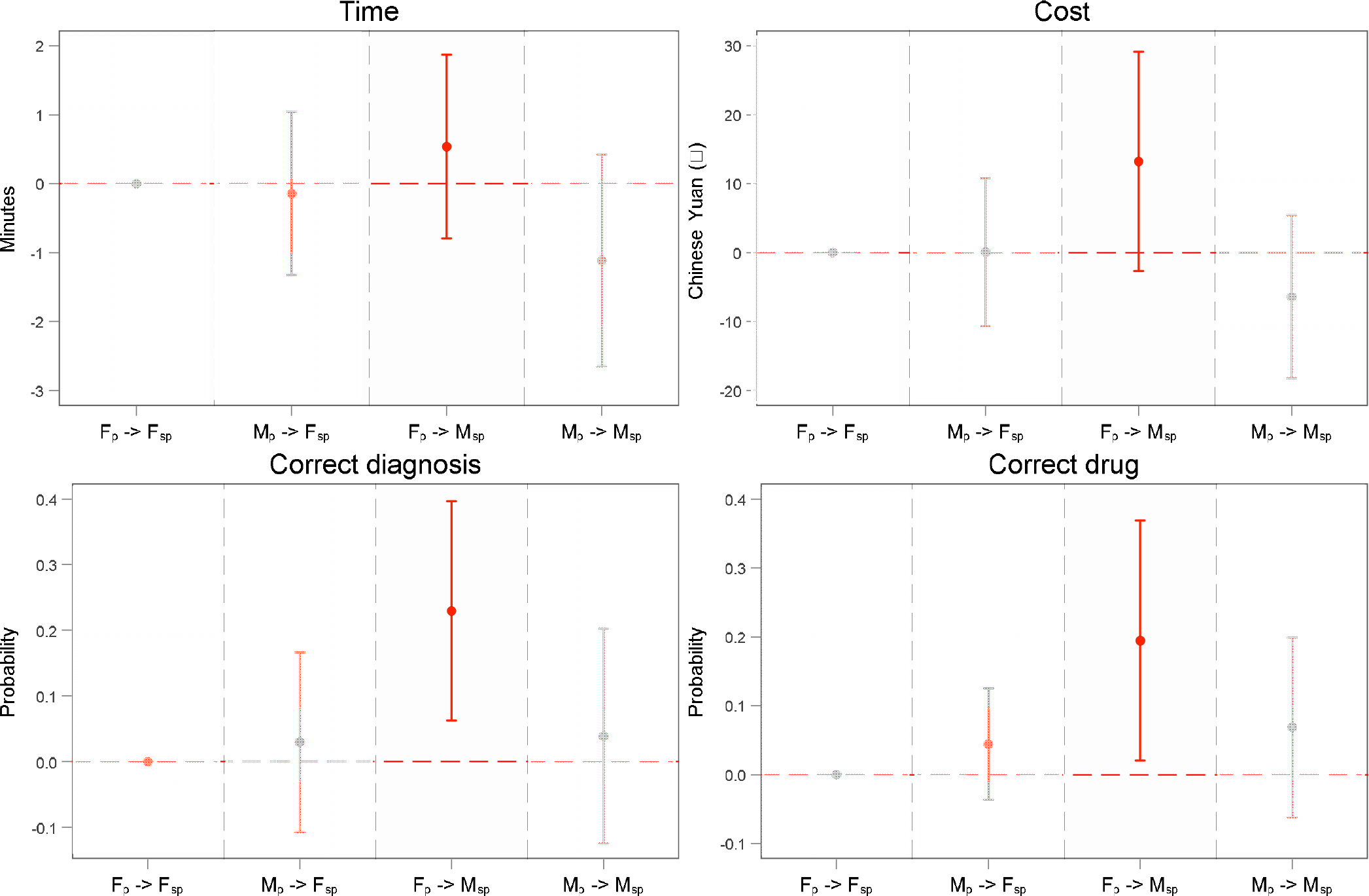
The impact of physician–patient gender match on healthcare quality Note: The pair of *F*_*p*_ -> *F* _*sp*_ denotes female physicians treating female patients (SPs) and others likewise. The figure was obtained by running our econometric specification. Physician age, CHC fixed effects, district fixed effects, disease fixed effects, month, day of the week, and year fixed effects were controlled for in the regressions. Robust standard errors, clustered at the CHC level. The details are reported in *Table S3*.

We investigate two channels to explain the gains in healthcare quality. First, physicians basically can do better in giving a correct diagnosis or drug prescriptions with more clinical information. However, we find no change in the adherence to a checklist of, either essential or recommended, guideline items (*Figure 3 Panel A*). Second, physician can communicate with patients more efficiently to achieve better performance. We find a 1.559 increase in patient-centered communications to find a common ground, although there was no significant change in the other two components (*Figure 3 Panel B*). Third, the gaps in healthcare quality were partly closed by patient-centered communications (*Table S6*) rather than by adherence to checklist items (*Table S7*). Fourth, since the lenient definition of healthcare quality leads to no penalty to unnecessary or even harmful tests and drug prescriptions, our additional investigations suggest that female physicians treating male SPs prescribed 0.623 more unnecessary tests and 0.299 fewer unnecessary drugs (*Figure 3 Panel C*).

**Figure 3.**
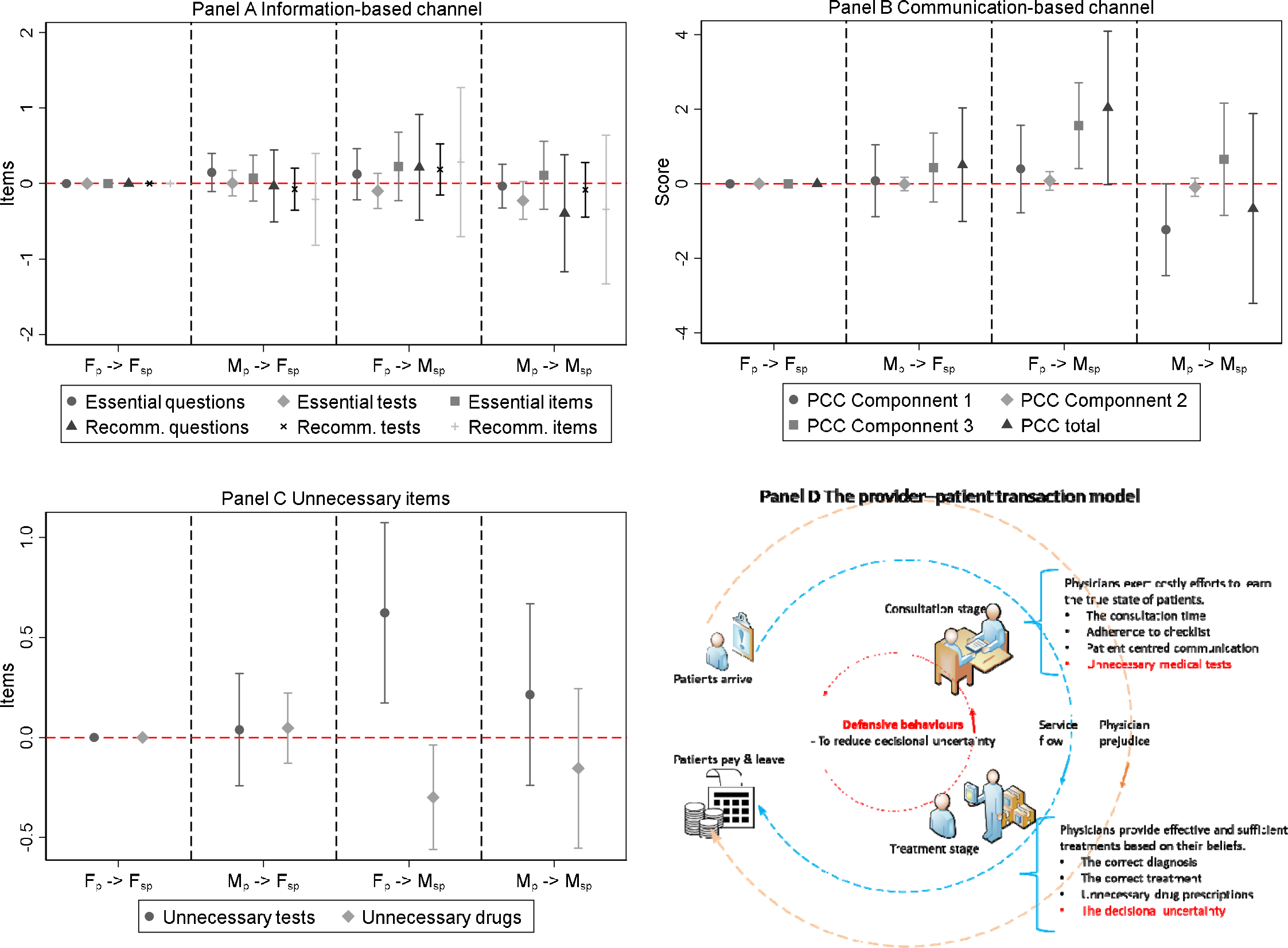
Explaining the gains in healthcare quality. Note: For Panel A-C, the pair of *F*_*p*_ -> *F* _*sp*_ F denotes female physicians treating female patients (SPs) and others likewise. Panel A-C were obtained by running our econometric specification. Physician age, CHC fixed effects, district fixed effects, disease fixed effects, month, day of the week, and year fixed effects were controlled for in the regressions. Robust standard errors, clustered at the CHC level. Their numeric values are reported in *Table S4, Table S5*, and *Table S8*. a) *Panel A:* Adherence to the checklist represents physicians’ adherence to a recommended checklist of inquiries and tests, serving as the benchmark for issuing a correct diagnosis and drug prescription (Das et al., 2012; Sylvia et al., 2015). It is a case-specific measure using count variables (see *Supplement 1*). b) *Panel B:* Patient-centered communication (PCC) represents a central clinical function (Stewart et al., 2013). We use three continuous variables in the study. Component 1 (exploring both the disease and the illness experience) refers to exploring physical or mental disorders of patients by history taking, physical, and laboratory examinations, and understanding a patient’s expectations and feelings. Component 2 (understanding the whole person) refers to understanding a patient’s disease and illness in the context of their life settings, including family, job, and social networks. Component 3 (finding common ground) refers to an agreement between physician and patient on three areas: the nature of the problem, the goals of treatment and disease management, and the roles of patient and physician. More details can be found in Su et al., (2022). c) *Panel C:* Unnecessary tests and drugs count the number of unnecessary medical tests and drugs prescribed for a specific interaction. A panel of physicians, pharmacists, and professors determined all medical tests and drugs in the study to be correct/essential, unnecessary, or harmful (see *Supplement 2*). d) *Panel D:* A provider’s job is to diagnose the true state of the patient and provide treatments accordingly (Das et al. 2016; Si et al. 2023). During the consultation stage, the provider exerts costly effort to learn about the true state of the patient through several checklist items, the time spent with the patient, and the medical tests prescribed. During the treatment stage, the provider determines the treatment types that they will provide based on their prior belief about the true state of the disease. The optimal outcome for a patient is to receive only the correct treatment, with no additional unnecessary treatments. In practice, providers exert the amount of effort and choose the treatments that will maximize their own utility, but these may not be aligned with those of the patient.

We perform five theory-based sensitivity analyses to confirm the robustness of our results. First, to rule out the possibility that our findings are being driven by a few SPs being particularly good actors in certain interactions, we perform falsification tests by randomly permuting observations in the treatment group. After running 500 repeated regressions, we find that the point estimates follow normal distributions (*Figure S1*), which suggests that no unexpected shocks were introduced. Second, our findings remain consistent when testing separately in the subsamples of female and male physicians (*Table S9*). Third, physician performance remains consistent when we introduce patient fixed effects (*Table S10*), suggesting the SPs were highly comparable. Fourth, in response to the literature that physician–patient gender concordance may improve healthcare quality, we further test this hypothesis, but our results do not lend support (*Table S11*). Fifth, we test the effect of age concordance (i.e., being in the same age bin of 40–50) between physicians and patients and find no impact (*Table S12*).

## Discussion

This experimental study shows that, compared with female physicians treating female patients, female physicians treating male patients provide a higher quality of care, including a 23.0 percentage-point increase in correct diagnosis and a 19.4 percentage-point increase in correct drug prescriptions. These disparities are much higher than the findings from equivalent studies in India, where the intensive training of informal healthcare providers was associated with only a 7.9 percentage-point increase in correct diagnoses and drug prescriptions (Das et al., 2016a). The differences in healthcare quality for female-female versus female-male pairings in our setting are very large, and, more importantly, we do not find any significant increase in overall medical costs and time investment for female physicians treating male patients.

Our findings in relation to healthcare quality are highly consistent with a bulk of literature finding that women face prevalent gender disparities and disadvantages in healthcare use and health outcomes (Clarke et al., 1994; Hamberg, 2008; Hoffmann and Tarzian, 2001; Milcent et al., 2007; Shannon et al., 2019). Our analyses suggest that the gains in healthcare quality were mainly attributed to better physician-patient communications, but not the presence of more clinical information. One plausible explanation for the identified impact is that physicians take greater care in their treatment of men because they are often the main income earners in families in China and worldwide (Chen and Ge, 2018; Chen et al., 2013). Therefore, male patients may receive more serious consideration than their female counterparts during a medical consultation, leading to an overall higher probability of correct diagnosis and drug prescriptions. However, this hypothesis fails to explain why we only see the impact when female physicians treat male SPs but not when male physicians treat male SPs.

The second potential explanation is that female physicians, in an attempt to practice defensive medicine, are more cautious in their diagnosis and treatment of male patients. Previous studies have found that physicians often respond to risks of malpractice litigation and workplace violence (Avraham and Schanzenbach, 2015; Currie and MacLeod, 2008; Frakes and Gruber, 2019; He, 2014; Keane et al., 2020; Kessler and McClellan, 1996) by rejecting high-risk patients, conducting fewer surgeries, performing more diagnostic tests, and prescribing more conservative treatment (Jia et al., 2021). In China, 62 percent of physicians reported experiencing workplace violence in 2017. This high rate persists, with physicians working in primary care settings experiencing the highest rates of serious workplace violence (Bo et al., 2020; Cai et al., 2019; Xu, 2014). In such cases, female physicians have an overwhelmingly higher risk of being the victims than their male counterparts do, while the perpetrators are usually men, few of whom have any criminal record or diagnosed mental illness (Hesketh et al., 2012).

Female physicians thus have a stronger motivation to defend themselves when treating male patients and may try harder to mitigate the risk of misdiagnosis (Ouyang, 2022). For example, female physicians exert more effort during the consultation stage to learn the true state of a disease when treating male patients, even though such effort can sometimes be clinically unnecessary. However, female physicians prescribe fewer drugs at the treatment stage to ensure that the total medical costs for male patients remain at an easily comparable level with that of other patients. In addition, the consultation time for female physicians treating male physicians was similar to the time they spent treating female patients, indicating that female physicians are shifting their efforts from the treatment stage to the consultation stage. This is clearly explained a two-stage transaction model (*Figure 3, Panel D*).

Our SP study design mitigates concerns over patient sorting within and across clinics, and the standardized set of patients and conditions also makes physicians’ performance highly comparable. However, we acknowledge three limitations. First, only a few diseases qualify for the SP approach. Although a growing number of studies implement the SP approach, its generalization deserves further research. However, a previous study has suggested that the patterns observed for the SP method and the patterns observed for real patients are essentially very similar (Das et al., 2016b). Second, more female SPs than male SPs were successfully recruited, which can be partially explained by greater opportunity costs for males to participate in the study than females. Further SP research should therefore seek to achieve a better SP gender balance in their recruitment. Third, to mitigate the concern that physicians’ exposure to medical violence may confound or exaggerate our findings, we conduct a systematic review using *China Core Newspaper*’s Full-Text Database. At least in our study setting and study period in Xi’an during the period 1998–2018, we identified no event of severe violence against medical professionals (i.e., physical injury or murder), which would have provided a strong incentive to physicians to change their behavior.

This study deepens our understanding of physicians’ decision-making process and sheds light on the potential to achieve higher-quality care in clinical practice through, for example, improving patient-centered communications and shifting effort from the treatment stage to the earlier stage of consultation, without creating excessive workloads for physicians and additional medical expenses for patients. Our findings also help us to understand the mechanism of physicians’ know-do gaps identified in medical practice (Leonard and Masatu, 2010; Mohanan et al., 2015) and the widely held notion that current health systems in many developing countries fail to motivate physicians to reach their productivity frontier in practice (Kovacs and Lagarde, 2022). For policy makers, compared with the traditional perception of using a clinical-training strategy, designing innovative interventions to improve patient centeredness and to incentivize physicians’ efforts in consultation can lead to substantial gains in the use of some inexpensive but potentially lifesaving diagnoses and treatments and equalize gender inequality in medical care.

## Data availability

The data are not publicly available due to restrictions of the ethics approval for this study. The code scripts used in this analysis are available from the corresponding authors upon reasonable request.

## Funding

This study was funded by National Natural Science Foundation of China (71874137) and China Medical Board (15-227), ARC Centre of Excellence in Population Ageing Research (project CE170100005), University of New South Wales, the U.S. PEPPER Center Scholar Award (P30AG021342), and two NIH/NIA grants (R01AG077529; K01AG053408).

## Contributors

X Chen and Z Zhou lead the research. Y Si and M Su conducted the field survey, collected and analyzed the data. Y Si and X Chen participated in the study design, data analysis and interpretation, and were the primary persons responsible for drafting the manuscript. G Chen, M Su, Z Zhou, and W Yip contributed to revision. All authors read and approved the final manuscript.

## Acknowledgement

The authors are grateful to all standardized patients and student instructors for collecting the data. The data are not publicly available due to restrictions of ethic approval requirements for this study. The code scripts used in this analysis are available from the corresponding authors upon reasonable request. The paper has been presented at Xi’an Jiaotong University, Shanghai Jiao Tong University, CHPAMS 2019 conference, Sichuan University, Peking University, Nanjing Medical University, Jinan University, iHEA 2019 Congress, Jinan University, Southeast University, and the University of New South Wales. The authors are grateful to the participants of these seminars and conferences for their helpful comments.

## Declaration of interests

we declare no competing interests.

## Notes

### Competing Interest Statement

The authors have declared no competing interest.

### Author Declarations

The study was approved by the Ethics Committee of Xi'an Jiaotong University Health Science Center (approval number 2015-406). This committee also permitted the recording of interactions between physicians and SPs using hidden audio devices.

