## Supplementary information for "The Impact of Physician-Patient Gender Match on Healthcare Quality: An Experiment in China"

Supplement 1 Checklist and diagnosis

*1.1 Adherence to the Checklist*

For asthma, there were 20 question items and 5 examination items in the checklist, among which 5 items were considered essential. For unstable angina, there were 22 question items and 5 examination items in the checklist of unstable angina, among which 6 items were considered essential.

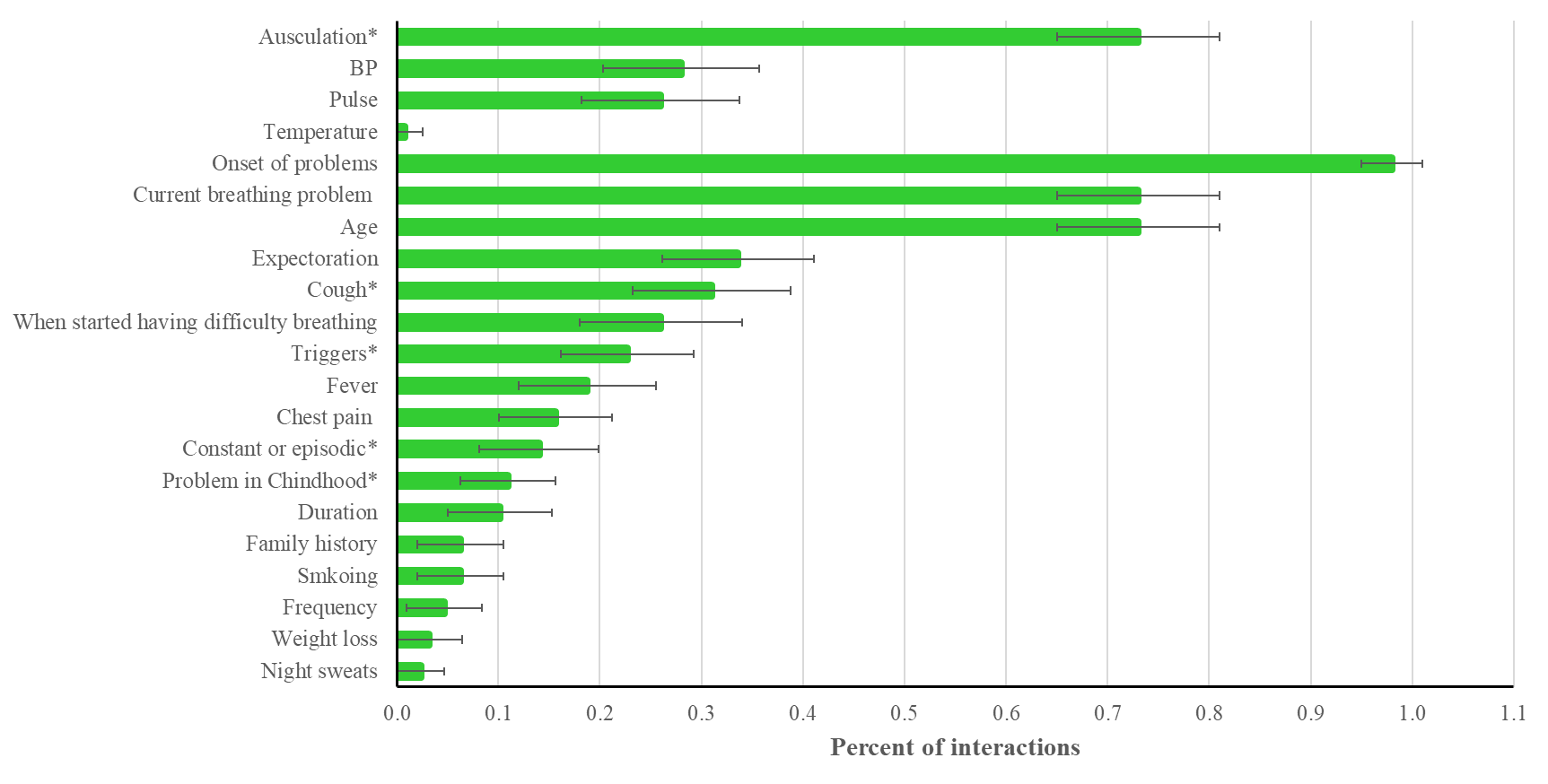

Figure 1.1.1 For asthma

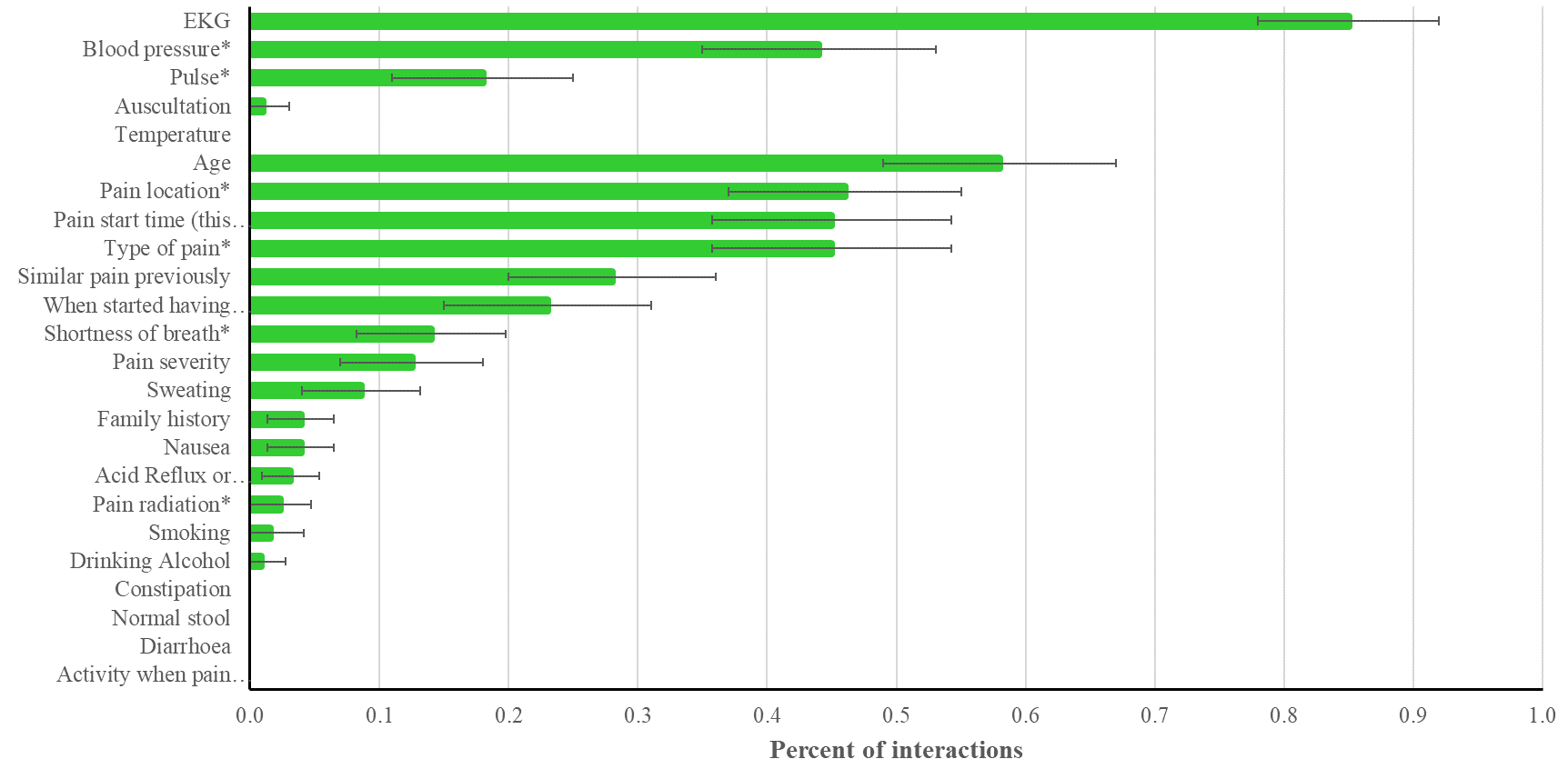

Figure 1.1.2 For unstable angina

Source: Author’s analysis.

Notes: Mean values and 95% confidence intervals are presented. All items are recommended and items with asterisks (*) regarded essential.

*1.2 Classification of Diagnoses for asthma*

Readers may contact the corresponding authors to request access to this Table/Figure.

### Supplement 2 Drug list and classifications

Readers may contact the corresponding authors to request access to this Table/Figure.

### Table S1 Balance check

| Matching | Public | Private | P-value | Non-  alliance | Alliance | P-value | Age  < 30 | 30 <=  Age <40 | 40 <=  Age <50 | Age  >= 50 | P-value |
| --- | --- | --- | --- | --- | --- | --- | --- | --- | --- | --- | --- |
| $F_{p} \& F_{sp}$ | 188 | 39 | 0.486 | 199 | 28 | 0.452 | 16 | 73 | 97 | 41 | <0.001 |
|  | 82.82 | 17.18 |  | 87.67 | 12.33 |  | 7.05 | 32.16 | 42.73 | 18.06 |  |
| $M_{p} \& F_{sp}$ | 155 | 29 |  | 167 | 17 |  | 7 | 31 | 52 | 94 |  |
|  | 84.24 | 15.76 |  | 90.76 | 9.24 |  | 3.8 | 16.85 | 28.26 | 51.09 |  |
| $F_{p} \& M_{sp}$ | 34 | 7 |  | 37 | 4 |  | 3 | 12 | 16 | 10 |  |
|  | 82.93 | 17.07 |  | 90.24 | 9.76 |  | 7.32 | 29.27 | 39.02 | 24.39 |  |
| $M_{p} \& M_{sp}$ | 37 | 3 |  | 33 | 7 |  | 2 | 8 | 15 | 15 |  |
|  | 92.5 | 7.5 |  | 82.5 | 17.5 |  | 5 | 20 | 37.5 | 37.5 |  |
| Total | 414 | 78 |  | 436 | 56 |  | 28 | 124 | 180 | 160 |  |
|  | 84.15 | 15.85 |  | 88.62 | 11.38 |  | 5.69 | 25.2 | 36.59 | 32.52 |  |

Note: The pair of $F_{p} \& F_{sp}$ denotes female physicians treating female patients (SPs) and others likewise. Chi-square test was used for dummy variable.

### Tabel S2 Quality metrics over physician–patient gender match

|  |  | $F_{p} \& F_{sp}$ | | $M_{p} \& F_{sp}$ | | $F_{p} \& M_{sp}$ | | $M_{p} \& M_{sp}$ | |  |
| --- | --- | --- | --- | --- | --- | --- | --- | --- | --- | --- |
|  |  | Mean | S.D. | Mean | S.D. | Mean | S.D. | Mean | S.D. | P-value |
| **Main outcomes** |  |  |  |  |  |  |  |  |  |  |
| Consultation length (Minutes) |  | 6.31 | 4.71 | 6.28 | 4.33 | 6.68 | 4.44 | 4.79 | 4.28 | 0.206 |
| Medical costs (CNY) |  | 33.0 | 41.1 | 35.8 | 39.7 | 51.8 | 53.6 | 25.3 | 29.7 | 0.022 |
| Correct diagnosis |  | 0.427 | 0.496 | 0.418 | 0.495 | 0.585 | 0.499 | 0.475 | 0.506 | 0.240 |
| Correct drug |  | 0.093 | 0.29 | 0.098 | 0.298 | 0.293 | 0.461 | 0.125 | 0.335 | 0.002 |
| **Information-based metrics** |  |  |  |  |  |  |  |  |  |  |
| Essential questions |  | 1.48 | 1.04 | 1.49 | 0.93 | 1.56 | 0.81 | 1.30 | 0.76 | 0.639 |
| Essential tests |  | 0.67 | 0.56 | 0.72 | 0.59 | 0.54 | 0.55 | 0.48 | 0.64 | 0.049 |
| Essential items (questions + tests) |  | 2.23 | 1.23 | 2.27 | 1.23 | 2.49 | 1.14 | 2.38 | 1.25 | 0.624 |
| Recommended questions |  | 4.93 | 2.79 | 4.68 | 2.58 | 6.00 | 2.73 | 4.68 | 2.23 | 0.037 |
| Recommended tests |  | 1.59 | 0.98 | 1.58 | 1.02 | 1.46 | 0.95 | 1.40 | 1.01 | 0.620 |
| Recommended items (questions + tests) |  | 7.07 | 3.21 | 6.84 | 3.27 | 8.39 | 3.41 | 7.38 | 3.45 | 0.050 |
| **Communication-based metrics** |  |  |  |  |  |  |  |  |  |  |
| Total score |  | 22.99 | 6.53 | 23.26 | 6.08 | 25.20 | 5.72 | 22.30 | 5.50 | 0.151 |
| Component 1 |  | 12.31 | 4.25 | 12.17 | 4.05 | 13.22 | 3.46 | 11.15 | 3.13 | 0.143 |
| Component 2 |  | 0.77 | 0.61 | 0.82 | 0.66 | 0.85 | 0.65 | 0.70 | 0.65 | 0.636 |
| Component 3 |  | 9.91 | 3.56 | 10.28 | 3.60 | 11.12 | 3.57 | 10.45 | 3.83 | 0.217 |
| **Unnecessary items** |  |  |  |  |  |  |  |  |  |  |
| Unnecessary tests |  | 0.87 | 0.98 | 0.81 | 1.05 | 1.54 | 1.25 | 0.95 | 1.06 | <0.001 |
| Unnecessary drugs |  | 0.46 | 0.82 | 0.52 | 0.83 | 0.17 | 0.59 | 0.33 | 0.94 | 0.075 |

Note: The pair of $F_{p} \& F_{sp}$ denotes female physicians treating female patients (SPs) and others likewise. CNY denotes Chinese yuan (exchange rate, 6.37 CNY ≈ 1 US dollar). Component1, Component 2 and Component 3 represent the three components of patient-centered communication, namely exploring both the disease and illness experience, understanding the whole person, and finding common ground. F&M denotes female physicians treating male patients. The statistical differences were analyzed using the chi-square test for binary variables and analysis of variance for continuous variables. S.D. means standard deviation.

### Table S3 The impact of physician–patient gender match on healthcare quality

|  | Time |  | Costs | Correct diagnosis | Correct drug |
| --- | --- | --- | --- | --- | --- |
| *Match* |  |  |  |  |  |
| $F_{p} \& F_{sp}$ | Ref. |  | Ref. | Ref. | Ref. |
| $M_{p} \& F_{sp}$ | -0.144 |  | 0.068 | 0.030 | 0.044 |
|  | (0.592) |  | (5.376) | (0.069) | (0.041) |
| $F_{p} \& M_{sp}$ | 0.537 |  | 13.220 | 0.230*** | 0.194** |
|  | (0.668) |  | (7.961) | (0.084) | (0.087) |
| $M_{p} \& M_{sp}$ | -1.116 |  | -6.411 | 0.039 | 0.069 |
|  | (0.770) |  | (5.935) | (0.082) | (0.065) |
| *N* | 492 |  | 492 | 492 | 492 |
| *R*^2^ | 0.275 |  | 0.375 | 0.342 | 0.185 |

Note: The pair of $F_{p} \& F_{sp}$ denotes female physicians treating female patients (SPs) and others likewise. The table was obtained by running our econometric specification. Physician age, CHC fixed effects, district fixed effects, disease fixed effects, month, day of the week, and year fixed effects were controlled for in the regressions. Robust standard errors, clustered at the CHC level, are presented in parentheses. *10% significance level. **5% significance level. ***1% significance level.

### Tabel S4 Essential items in the checklist

|  | (1) | (2) | (3) | (4) | (5) | (6) |
| --- | --- | --- | --- | --- | --- | --- |
|  | Essential questions | Essential  tests | Essential  items | Recommened questions | Recommened  tests | recommended items |
| $F_{p} \& F_{sp}$ | Ref. | Ref. | Ref. | Ref. | Ref. | Ref. |
| $M_{p} \& F_{sp}$ | 0.147 | 0.00579 | 0.0719 | -0.0326 | -0.0746 | -0.210 |
|  | (0.126) | (0.0841) | (0.151) | (0.238) | (0.139) | (0.305) |
| $F_{p} \& M_{sp}$ | 0.124 | -0.0994 | 0.225 | 0.216 | 0.186 | 0.284 |
|  | (0.169) | (0.116) | (0.228) | (0.350) | (0.169) | (0.495) |
| $M_{p} \& M_{sp}$ | -0.0332 | -0.226^*^ | 0.109 | -0.395 | -0.0830 | -0.343 |
|  | (0.146) | (0.125) | (0.225) | (0.389) | (0.182) | (0.493) |
| *N* | 492 | 492 | 492 | 492 | 492 | 492 |
| *R*^2^ | 0.222 | 0.253 | 0.268 | 0.527 | 0.435 | 0.505 |

Note: The pair of $F_{p} \& F_{sp}$ denotes female physicians treating female patients (SPs) and others likewise. The table was obtained by running our econometric specification. Physician age, CHC fixed effects, district fixed effects, disease fixed effects, month, day of the week, and year fixed effects were controlled for in the regressions. Robust standard errors, clustered at the CHC level, are presented in parentheses. *10% significance level. **5% significance level. ***1% significance level.

### Table S5 Patient-centered communications

|  | (1) | (2) | (3) | (4) |
| --- | --- | --- | --- | --- |
|  | PCC  Component 1 | PCC  Component 1 | PCC  Component 1 | PCC  Total score |
| $F_{p} \& F_{sp}$ | Ref. | Ref. | Ref. | Ref. |
| $M_{p} \& F_{sp}$ | 0.0809 | -0.00618 | 0.434 | 0.509 |
|  | (0.485) | (0.0887) | (0.461) | (0.763) |
| $F_{p} \& M_{sp}$ | 0.401 | 0.0799 | 1.559^***^ | 2.040^*^ |
|  | (0.587) | (0.125) | (0.577) | (1.030) |
| $M_{p} \& M_{sp}$ | -1.231^*^ | -0.0936 | 0.658 | -0.666 |
|  | (0.616) | (0.123) | (0.755) | (1.273) |
| *N* | 492 | 492 | 492 | 492 |
| *R*^2^ | 0.405 | 0.273 | 0.268 | 0.310 |

Note: The pair of $F_{p} \& F_{sp}$ denotes female physicians treating female patients (SPs) and others likewise. The table was obtained by running our econometric specification. Physician age, CHC fixed effects, district fixed effects, disease fixed effects, month, day of the week, and year fixed effects were controlled for in the regressions. PCC C1, C2, and C3 represent the three components of patient-centered communication, namely exploring both the disease and illness experience, understanding the whole person, and finding common ground. Robust standard errors, clustered at the CHC level, are presented in parentheses. *10% significance level. **5% significance level. ***1% significance level.

### Table S6 Mediation analyses via communication-based channel

|  | (1) | (2) | (3) | (4) |
| --- | --- | --- | --- | --- |
|  | Correct diagnosis | Correct drug | Correct diagnosis | Correct drug |
| Gender match |  |  |  |  |
| $F_{p} \& F_{sp}$ | Ref. | Ref. | Ref. | Ref. |
| $M_{p} \& F_{sp}$ | 0.0223 | 0.0374 | 0.0136 | 0.0354 |
|  | (0.0663) | (0.0373) | (0.0621) | (0.0369) |
| $F_{p} \& M_{sp}$ | 0.200^**^ | 0.167^*^ | 0.176^**^ | 0.158^*^ |
|  | (0.0834) | (0.0843) | (0.0772) | (0.0843) |
| $M_{p} \& M_{sp}$ | 0.0485 | 0.0777 | 0.00666 | 0.0657 |
|  | (0.0835) | (0.0635) | (0.0837) | (0.0628) |
| PCC total score | 0.0147^***^ | 0.0134^***^ |  |  |
|  | (0.00496) | (0.00282) |  |  |
| PCC Component 1 |  |  | -0.00328 | 0.00530 |
|  |  |  | (0.00744) | (0.00523) |
| PCC Component 2 |  |  | -0.0371 | 0.0370 |
|  |  |  | (0.0412) | (0.0365) |
| PCC Component 3 |  |  | 0.0374^***^ | 0.0199^***^ |
|  |  |  | (0.00646) | (0.00501) |
| *N* | 492 | 492 | 492 | 492 |
| *R*^2^ | 0.366 | 0.233 | 0.393 | 0.240 |

Note: The pair of $F_{p} \& F_{sp}$ denotes female physicians treating female patients (SPs) and others likewise. The table was obtained by running our econometric specification. Physician age, CHC fixed effects, district fixed effects, disease fixed effects, month, day of the week, and year fixed effects were controlled for in the regressions. Robust standard errors, clustered at the CHC level, are presented in parentheses. *10% significance level. **5% significance level. ***1% significance level.

### Table S7 Mediation analyses via information-based channel

|  | (1) | (2) | (3) | (4) |
| --- | --- | --- | --- | --- |
|  | Correct diagnosis | Correct drug | Correct diagnosis | Correct drug |
| Gender match |  |  |  |  |
| $F_{p} \& F_{sp}$ | Ref. | Ref. | Ref. | Ref. |
| $M_{p} \& F_{sp}$ | 0.0301 | 0.0473 | 0.0302 | 0.0449 |
|  | (0.0685) | (0.0394) | (0.0693) | (0.0405) |
| $F_{p} \& M_{sp}$ | 0.229^***^ | 0.190^**^ | 0.228^***^ | 0.190^**^ |
|  | (0.0838) | (0.0877) | (0.0842) | (0.0863) |
| $M_{p} \& M_{sp}$ | 0.0394 | 0.0738 | 0.0413 | 0.0760 |
|  | (0.0816) | (0.0640) | (0.0817) | (0.0643) |
| Recommended items | 0.00185 | 0.0146^*^ |  |  |
|  | (0.0115) | (0.00768) |  |  |
| Recommended questions |  |  | 0.00569 | 0.0181 |
|  |  |  | (0.0138) | (0.0115) |
| Recommended tests |  |  | 0.00344 | 0.00112 |
|  |  |  | (0.0330) | (0.0182) |
| *N* | 492 | 492 | 492 | 492 |
| *R*^2^ | 0.342 | 0.196 | 0.343 | 0.196 |

Note: The pair of $F_{p} \& F_{sp}$ denotes female physicians treating female patients (SPs) and others likewise. The table was obtained by running our econometric specification. Physician age, CHC fixed effects, district fixed effects, disease fixed effects, month, day of the week, and year fixed effects were controlled for in the regressions. Robust standard errors, clustered at the CHC level, are presented in parentheses. *10% significance level. **5% significance level. ***1% significance level.

### Table S8 Unnecessary items

|  | Unnecessary tests |  | Unnecessary drugs |
| --- | --- | --- | --- |
| *Match* |  |  |  |
| $F_{p} \& F_{sp}$ | Ref. |  | Ref. |
| $M_{p} \& F_{sp}$ | 0.039 |  | 0.047 |
|  | (0.140) |  | (0.088) |
| $F_{p} \& M_{sp}$ | 0.623*** |  | -0.299** |
|  | (0.225) |  | (0.131) |
| $M_{p} \& M_{sp}$ | 0.214 |  | -0.154 |
|  | (0.227) |  | (0.199) |
| *N* | 492 |  | 492 |
| *R*^2^ | 0.353 |  | 0.291 |

Note: The pair of $F_{p} \& F_{sp}$ denotes female physicians treating female patients (SPs) and others likewise. The table was obtained by running our econometric specification. Physician age, CHC fixed effects, district fixed effects, disease fixed effects, month, day of the week, and year fixed effects were controlled for in the regressions. Robust standard errors, clustered at the CHC level, are presented in parentheses. *10% significance level. **5% significance level. ***1% significance level.

### Figure S1 Falsification test

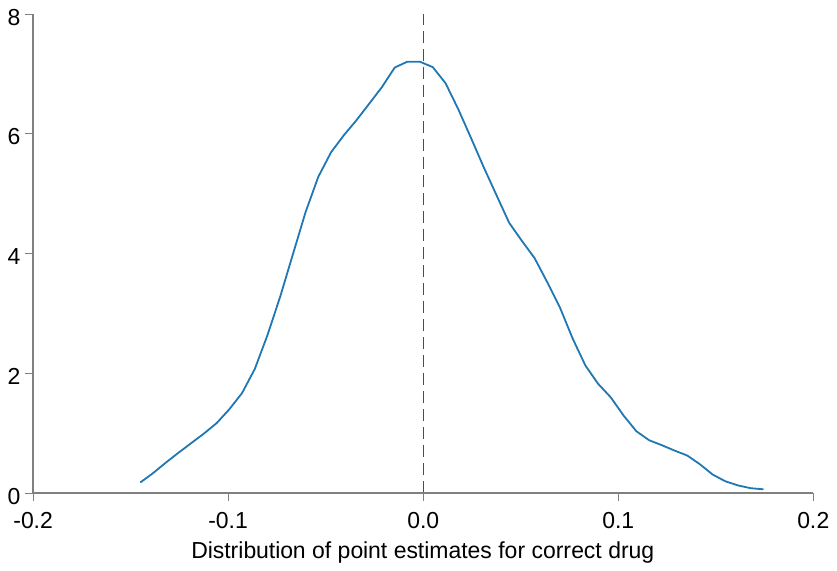

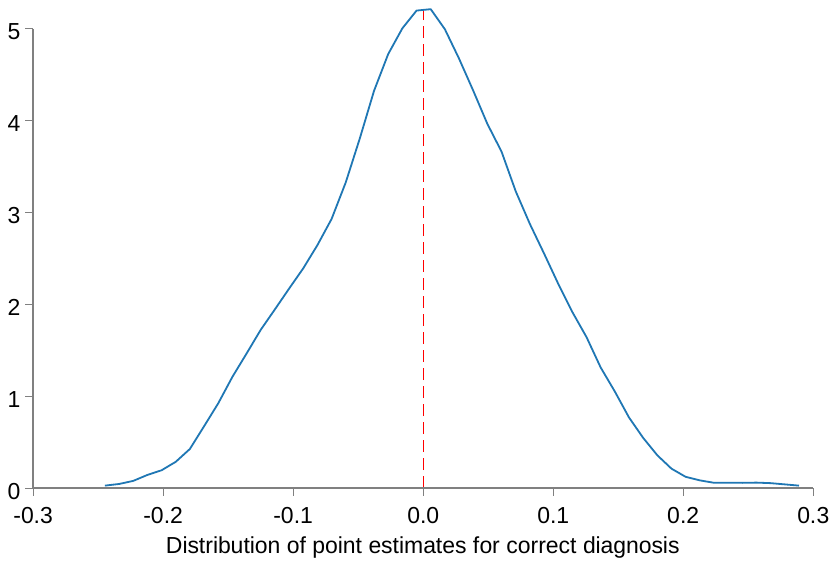

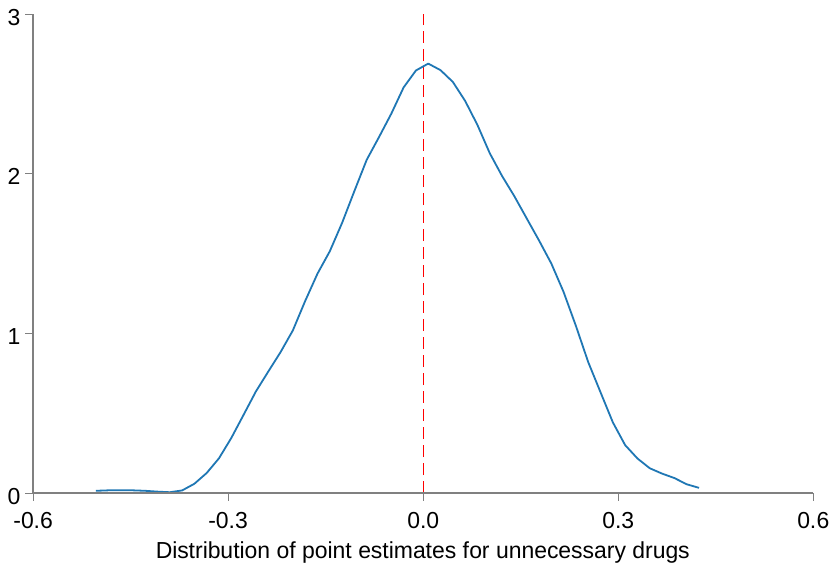

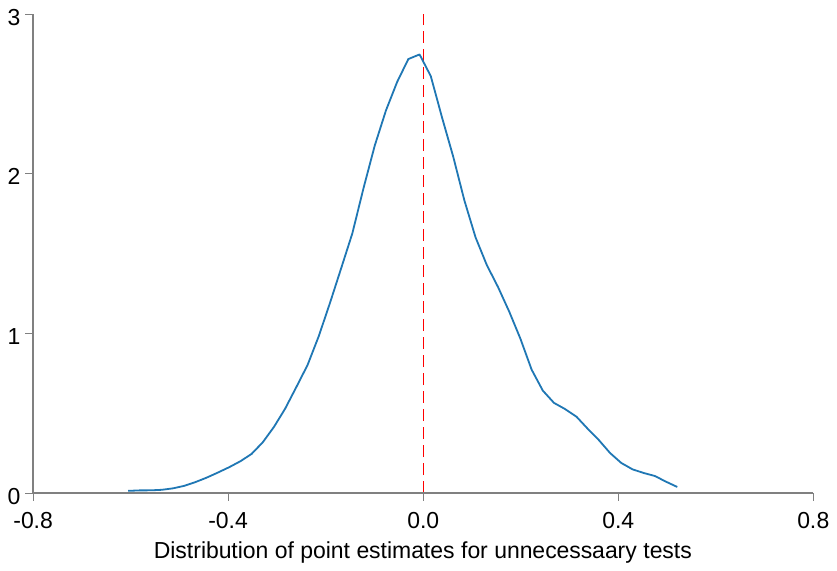

Note: In the analysis, 500 repeated regressions were estimated and here we plot the distribution of coefficients for female physicians treating male SPs. The coefficients were obtained by running our econometric specification. Physician age, CHC fixed effects, district fixed effects, disease fixed effects, month, day of the week, and year fixed effects were controlled for in the regressions. Robust standard errors, clustered at the CHC level, are presented in parentheses.

### Table S9 Subsample estimates by physician gender

*Subsample of female physicians*

|  | (1) | (2) | (3) | (4) | (5) | (6) | (7) | (8) | (9) | (10) |
| --- | --- | --- | --- | --- | --- | --- | --- | --- | --- | --- |
|  | Q+E | Consultation  time | Correct  diagnosis | Correct  drug | Unnecessary  tests | Unnecessary  drugs | Total  cost | C1 | C2 | C3 |
| Male SP | -0.150 | 0.622 | 0.168* | 0.168* | 0.623** | -0.364** | 12.53 | -0.314 | 0.0671 | 1.310* |
|  | (0.543) | (0.796) | (0.0909) | (0.0995) | (0.255) | (0.136) | (9.028) | (0.633) | (0.138) | (0.678) |
| County fixed effect | Yes | Yes | Yes | Yes | Yes | Yes | Yes | Yes | Yes | Yes |
| CHC fixed effect | Yes | Yes | Yes | Yes | Yes | Yes | Yes | Yes | Yes | Yes |
| Day of week, year fixed effect | Yes | Yes | Yes | Yes | Yes | Yes | Yes | Yes | Yes | Yes |
| Case fixed effect | Yes | Yes | Yes | Yes | Yes | Yes | Yes | Yes | Yes | Yes |
| *N* | 268 | 268 | 268 | 268 | 268 | 268 | 268 | 268 | 268 | 268 |
| *R*^2^ | 0.596 | 0.365 | 0.447 | 0.291 | 0.532 | 0.342 | 0.472 | 0.508 | 0.407 | 0.391 |

Note: Table reports OLS estimates of Specification 1 with controls for physician age. CHC fixed effects, district fixed effects, disease fixed effects, month, day of the week, and year fixed effects were controlled for in the regressions. Q represents adherence to recommended questions; E represents adherence to recommended tests; C1, C2 and C3 represent the three components of patient-centered communication, namely exploring both the disease and illness experience, understanding the whole person, and finding common ground. Robust standard errors, clustered at the community health center level, are presented in parentheses. *10% significance level. **5% significance level. ***1% significance level.

*Subsample of male physicians*

|  | (1) | (2) | (3) | (4) | (5) | (6) | (7) | (8) | (9) | (10) |
| --- | --- | --- | --- | --- | --- | --- | --- | --- | --- | --- |
|  | Q+E | Consultation  time | Correct  diagnosis | Correct  drug | Unnecessary  tests | Unnecessary  drugs | Total  cost | C1 | C2 | C3 |
| Male SP | 0.00464 | -0.882 | 0.0241 | 0.0538 | 0.262 | -0.177 | -8.512 | -0.858 | -0.00826 | 0.165 |
|  | (0.601) | (0.878) | (0.104) | (0.0666) | (0.192) | (0.219) | (7.074) | (0.744) | (0.124) | (0.851) |
| County fixed effect | Yes | Yes | Yes | Yes | Yes | Yes | Yes | Yes | Yes | Yes |
| CHC fixed effect | Yes | Yes | Yes | Yes | Yes | Yes | Yes | Yes | Yes | Yes |
| Day of week, year fixed effect | Yes | Yes | Yes | Yes | Yes | Yes | Yes | Yes | Yes | Yes |
| Case fixed effect | Yes | Yes | Yes | Yes | Yes | Yes | Yes | Yes | Yes | Yes |
| *N* | 224 | 224 | 224 | 224 | 224 | 224 | 224 | 224 | 224 | 224 |
| *R*^2^ | 0.568 | 0.533 | 0.575 | 0.331 | 0.454 | 0.502 | 0.466 | 0.521 | 0.464 | 0.353 |

Note: Table reports OLS estimates of Specification 1 with controls for physician age. CHC fixed effects, district fixed effects, disease fixed effects, month, day of the week, and year fixed effects were controlled for in the regressions. Q represents adherence to recommended questions; E represents adherence to recommended tests; C1, C2 and C3 represent the three components of patient-centered communication, namely exploring both the disease and illness experience, understanding the whole person, and finding common ground. Robust standard errors, clustered at the community health center level, are presented in parentheses. *10% significance level. **5% significance level. ***1% significance level.

### Table S10 Physician performance with patient fixed effects

|  | (1) | (2) | (3) | (4) | (5) | (6) | (7) | (8) | (9) | (10) |
| --- | --- | --- | --- | --- | --- | --- | --- | --- | --- | --- |
|  | Q+E | Consultation  time | Correct  diagnosis | Correct  drug | Unnecessary  tests | Unnecessary  drugs | Total  cost | C1 | C2 | C3 |
| Female physician | -0.259 | -0.469 | -0.0260 | 0.00868 | -0.00793 | 0.0189 | -4.308 | -0.205 | -0.0639 | 0.0399 |
|  | (0.289) | (0.568) | (0.0646) | (0.0401) | (0.127) | (0.0904) | (4.697) | (0.458) | (0.0713) | (0.360) |
| *N* | 492 | 492 | 492 | 492 | 492 | 492 | 492 | 492 | 492 | 492 |
| *R*^2^ | 0.661 | 0.421 | 0.365 | 0.210 | 0.387 | 0.371 | 0.408 | 0.518 | 0.344 | 0.454 |

Note: Table reports OLS estimates of Specification 1 with controls for physician age. CHC fixed effects, district fixed effects, disease fixed effects, month, day of the week, and year fixed effects were controlled for in the regressions. Q represents adherence to recommended questions; E represents adherence to recommended tests; C1, C2 and C3 represent the three components of patient-centered communication, namely exploring both the disease and illness experience, understanding the whole person, and finding common ground. Robust standard errors, clustered at the community health center level, are presented in parentheses. *10% significance level. **5% significance level. ***1% significance level.

### Table S11 Physician-patient gender concordance

|  | (1) | (2) | (3) | (4) | (5) | (6) | (7) | (8) | (9) | (10) |
| --- | --- | --- | --- | --- | --- | --- | --- | --- | --- | --- |
|  | Q+E | Consultation  time | Correct  diagnosis | Correct  drug | Unnecessary  tests | Unnecessary  drugs | Total  cost | C1 | C2 | C3 |
| *Physician-patient gender concordance* | | | |  |  |  |  |  |  |  |
| Gender concordance | 0.002 | -0.286 | -0.075 | -0.069* | -0.147 | 0.011 | -5.003 | -0.439 | -0.038 | -0.589 |
|  | (0.270) | (0.450) | (0.0531) | (0.040) | (0.123) | (0.078) | (4.503) | (0.408) | (0.072) | (0.409) |
| *N* | 492 | 492 | 492 | 492 | 492 | 492 | 492 | 492 | 492 | 492 |
| *R*^2^ | 0.503 | 0.271 | 0.333 | 0.170 | 0.334 | 0.279 | 0.369 | 0.400 | 0.271 | 0.261 |

Note: Table reports OLS estimates of Specification 1 with controls for physician age. CHC fixed effects, district fixed effects, disease fixed effects, month, day of the week, and year fixed effects were controlled for in the regressions. Q represents adherence to recommended questions; E represents adherence to recommended tests; C1, C2 and C3 represent the three components of patient-centered communication, namely exploring both the disease and illness experience, understanding the whole person, and finding common ground. Robust standard errors, clustered at the community health center level, are presented in parentheses. *10% significance level. **5% significance level. ***1% significance level.

### Table S12 Physician-patient age concordance

|  | (1) | (2) | (3) | (4) | (5) | (6) | (7) | (8) | (9) | (10) |
| --- | --- | --- | --- | --- | --- | --- | --- | --- | --- | --- |
|  | Q+E | Consultation  time | Correct  diagnosis | Correct  drug | Unnecessary  tests | Unnecessary  drugs | Total  cost | C1 | C2 | C3 |
| Age  concordance | 0.172 | 0.743 | -0.0266 | -0.0217 | 0.0862 | 0.0435 | -1.139 | 0.0830 | 0.00362 | -0.635 |
|  | (0.298) | (0.513) | (0.0483) | (0.0321) | (0.113) | (0.0901) | (4.694) | (0.394) | (0.0689) | (0.395) |
| *N* | 492 | 492 | 492 | 492 | 492 | 492 | 492 | 492 | 492 | 492 |
| *R*^2^ | 0.502 | 0.270 | 0.332 | 0.170 | 0.345 | 0.288 | 0.358 | 0.397 | 0.266 | 0.266 |

Note: Table reports OLS estimates of Specification 1 with controls for physician gender and patient gender. CHC fixed effects, district fixed effects, disease fixed effects, month, day of the week, and year fixed effects were controlled for in the regressions. Q represents adherence to recommended questions; E represents adherence to recommended tests; C1, C2 and C3 represent the three components of patient-centered communication, namely exploring both the disease and illness experience, understanding the whole person, and finding common ground. Robust standard errors, clustered at the community health center level, are presented in parentheses. *10% significance level. **5% significance level. ***1% significance level.
